# Understanding Definitions and Reporting of Deaths Attributed to COVID-19 in the UK – Evidence from FOI Requests

**DOI:** 10.1101/2022.04.28.22274344

**Authors:** Tom Jefferson, Madeleine Dietrich, Jon Brassey, Carl Heneghan

## Abstract

Death is a widely used outcome to assess the severity of pandemics. Accuracy in assigning the cause of death is of vital importance to define the impact of the agent, monitor its evolution, and compare its threat with those of other agents. Throughout the COVID-19 pandemic, there has been widespread reporting of aggregate death data with little attention paid to the accuracy of the assignment of causation.

We aimed to analyse public authorities’ understanding of the assignment of cause of deaths during the SARS-CoV-2 pandemic in the UK by accessing Freedom of Information requests posed in three periods in 2020-21. By public authorities, we mean NHS Health Trusts, laboratories, and government agencies such as Public Health England and the Department of Health and Social Care. We searched WhatDoTheyKnow using the terms “covid and death”. We excluded those requests to bodies that cannot provide an answer (e.g. Councils) and those dealing with the effects of vaccines.

We grouped questions into themes addressing the definitions and causes of death relevant to the pandemic. We looked at the responses to the questions of the definition of cause of death, the accuracy of the attribution, the role of other pre-existing pathologies and how these were reported and quantified.

We found 800 requests from over 90 individuals. There was no consistency in the definition of cause of death or contributory cause of death across national bodies and in different bodies within the same nation. Nursing home providers, as well as medical practitioners, can assign a cause of death according to the Care Quality Commission. Post-mortem examinations were uncommon, the ONS did not incorporate their results in the summary of deaths by cause during the pandemic period. The meaning of the words “test” or “swab” was never clarified by any of the respondents. In care homes in England 1,304 out of 17,264 COVID-19 (7.6%, range 0% to 63%) mentioned COVID-19 in the absence of contributory or other factors in the death certificate, making it impossible to ascertain a chain of causality. The inconsistencies already noted hinder the ascertainment of the role of each factor leading to death and the quantification of the importance of infection. Some responses indicate that SARS-CoV-2 negative individuals or those whose death was not caused by COVID-19 were classified as “COVID-19 deaths”. We found 14 different ways of attributing the causes of death mentioned by respondents.

The overall lack of consistency has confused the public and likely led to erroneous conclusions. We are unable to separate the effects on deaths of SARS-CoV-2 from those of human interventions. A coherent process based on consistent definitions across the devolved nations is required. Furthermore, to enhance the accuracy of causation in pandemics a subset of deaths should be verified using autopsies with full medical documentation.

## Introduction

The two years of the COVID-19 pandemic restrictions were primarily underpinned by concerns over rising cases, hospital admissions and deaths. Deaths remain the most troubling as a marker of the severity of the disease. However, throughout the pandemic, it has not been possible to determine who has died ‘from’ or ‘with’ COVID.

To answer this vital question, data availability has not been the problem - several platforms have reported global daily updates on the latest number of deaths. However, there has been a scarcity of critical thinking about what individuals labelled as ‘COVID deaths’ died from. For example, we have documented that some studies attributed mortality in nursing homes in the absence of testing or a medical diagnosis.^1^ The authors inferred the cause of death from the period in which the deaths occurred.

It is often problematic for clinicians to determine the direct cause of death or the underlying cause: none of the publicised daily death tolls differentiates between the two. The true impact of the SARS-CoV-2 virus is difficult to determine if we cannot interpret the actual cause of death. Also, misunderstanding can easily be misinterpreted and lead to underestimations or misleading statements about the effect on deaths. Attributable deaths are an important yardstick to monitor the trajectory of a pandemic.

Accounting for those that died from COVID-19 related illness is complex.^2^ We are aware of several methods that label a death as causative, leading to significant differences in the reported number of COVID-19 deaths. For example, The Office for National Statistics’ weekly death counts in England and Wales are based on mentions on the death certificate. Daily figures published by the Department of Health and Social Care differ because they report deaths occurring within 28 days of a positive test.^3^

At the beginning of the pandemic, Public Health England linked data on positive cases to the NHS central register of patients who died. This definition meant that a patient who tested positive would be counted as a COVID death even if they were run over by a bus several months later. This method <u>over-estimated the COVID-associated deaths.</u> To overcome this limitation, the UK daily COVID death counts on the <u>COVID-19 dashboard</u> were changed to report deaths within 28 days of a first positive laboratory-confirmed test.

The ONS method establishes the cause of death through death certificates. However, anyone who has completed a death certificate will acknowledge their constraints and the potential inaccuracies for assigning causation.

Some commentators consider inaccuracies are not a problem in the certification process.^4^ However, a review of their accuracy in an intensive care unit showed up to 10% were completed to a poor standard, and just over a half to a minimum standard.^5^ A further study showed 82% of death certificates contained one or more errors,^6^ and a substantial discrepancy has been shown between the diagnoses on death certificates compared with autopsy.^7^ For example, certificates from 433 autopsied hospital patients in Iceland matched against post-mortem examinations showed significant discrepancies in 50% of patients and incorrectly stated the immediate cause of death in 25%.^8^

This problem in the assignment of the cause of death has not escaped the notice of the public who, through the Freedom of Information Act (“FOIA”), have made requests to substantiate causation of those patients labelled as COVID deaths. A clear understanding of the different methods for defining causation is vital before analysing the substantial amount of data collected during the COVID-19 pandemic.

The FOIA 2000 allows for a public “right of access” to information held by public authorities in the UK. The website <u>WhatDoTheyKnow</u> facilitates FOI requests by forwarding requests to the appropriate authority and publishing the subsequent responses. We used a similar approach in a previous report in the role of PCR Testing in the UK During the SARS-CoV-2 pandemic.^9^

We similarly collated FOI responses to preliminary understand the current knowledge of the causes of COVID-19 deaths in the UK – how such causation is assigned amongst public bodies and to what extent definitions differ depending on the organisation. We also provide an interpretation of the various FOI responses and recommendations to improve the understanding of the cause of COVID-19 deaths.

## Methods

We set out to analyse public authorities’ understanding of how deaths from SARS-CoV-2 are measured in the UK by accessing Freedom of Information requests posed in 2020-21. By public authorities, we mean NHS Health Trusts, laboratories, and government agencies such as Public Health England and the Department of Health and Social Care. We searched *WhatDoTheyKnow* using the terms “covid and death” to understand the ascertainment and cause of deaths in the UK. We found 800 requests, from over 90 individuals.

We searched through FOI requests using a cascade approach, keeping only those that gave informative results. We excluded those requests to bodies that cannot provide an answer (e.g. Councils) and those dealing with the effects of vaccines. We recorded the example and the respondent type and excluded any requests on causes of death other than COVID-19 and explanatory requests based on baseline characteristics such as age, place of death, etc. We also excluded questions posed to bodies such as councils with no statutory duty to collect death data. We concentrated on 3 periods of the pandemic assessing FOI answers provided in the spring and late 2020 and 2021, and we grouped questions into themes addressing the definitions and causes of death relevant to the pandemic.

## Results

### 1. Definition of COVID-19 Death

Any emphasis of quoted sections is ours.

#### Office for National Statistics (ONS)

The ONS provides a <u>weekly return</u>^10^ on the number of deaths based on Medical Certificates of Cause of Death (MCCD). When someone dies, a medical practitioner writes the medical certificate based on the cause of death, which is then recorded at a local authority registration office. This information is then sent electronically to the ONS to produce statistics about causes of death.

> *‘Provisional counts of the number of deaths registered in England and Wales, by age, sex and region, in the latest weeks for which data are available. Includes the weekly figures available for deaths involving COVID-19 and provides 5-year averages for comparisons*.*’*

Three responses provide information on the ONS definitions used:

ONS in a response to an FOI request produced this answer on <u>21 Sept 2021</u>:

> *‘When we say that a death ‘****involved’ COVID-19***, *we mean that COVID-19 was mentioned anywhere on the death certificate, possibly along with other health conditions, not necessarily as the underlying cause of death. When we say that a death was ‘****due to****’ COVID-19, we mean that COVID-19 was the underlying cause of death, because it was* ***either the only health condition mentioned on the death certificate, or it was the one that started the train of events leading to death***.*’*

ONS here clearly indicates that **“due to”**, is either 1a (the only condition) or 1b or c (the events trigger) in the death certificate which is used “*to produce statistics about causes of death”*.

In a response dated <u>23 Dec 2020</u> ONS state:

> *‘We use the term ‘Due to COVID-19’ when referring to deaths where COVID-19 was recorded as the underlying cause of death. We use the term ‘involving COVID-19’ when referring to deaths that had the illness mentioned anywhere on the death certificate, whether as an underlying cause or not*.*’*

This is not the same wording as that of the previous answer as the explanation of **underlying** and **only** are missing, as the cause that started the train of events leading to death. In this version of the definition of “Directly leading to death” (1a in the death certificate) is missing. This is strange given that ONS has a dedicated FOI answering apparatus and one would expect identical answers to identical questions.

On another response on <u>28 Feb 2022</u> ONS state:

> *‘We use the term “****due to****” a cause of death (e*.*g. COVID-19) when referring* ***only*** *to deaths with that underlying cause of death. We use the term “****involving****” when referring to all deaths that had the cause mentioned anywhere on the death certificate, whether as an underlying cause or not. For more information on cause of death coding see our User Guide to Mortality Statistics*.*’*

This version is missing the immediate cause of death that could occur if you used the <u>21 Sept 2021</u> definition.

#### NHS England

NHS England and NHS Improvement publish the number of patients who died in hospital and tested positive for COVID-19, whether they had a pre-existing condition or whether COVID-19 was mentioned on the death certificate. This can be found in the <u>weekly file</u> on their website.^11^ These figures don’t state whether or not COVID-19 was the single cause of death. All deaths are recorded against the date of death rather than the date they were announced. Figures do not include deaths outside hospitals, such as in care homes.

##### Definition

Covid Daily Deaths. <u>28 April 2020</u>

> *‘From Tuesday 28 April 2020, NHS England and NHS Improvement started to report the number of patient deaths where there has been no COVID-19 positive test result, but where COVID-19 is documented as a* ***direct*** *or* ***underlying*** *cause of death on part 1 or part 2 of the death certification process. This change has been introduced for deaths that occurred on 24th April and subsequently are shown separately in the region data table only*.*’*

<u>ONS Guidance</u> for doctors completing Medical Certificates of Cause of Death (MCCD) in England and Wales *for use during the emergency period only* reports the MCCD is set out in two parts:

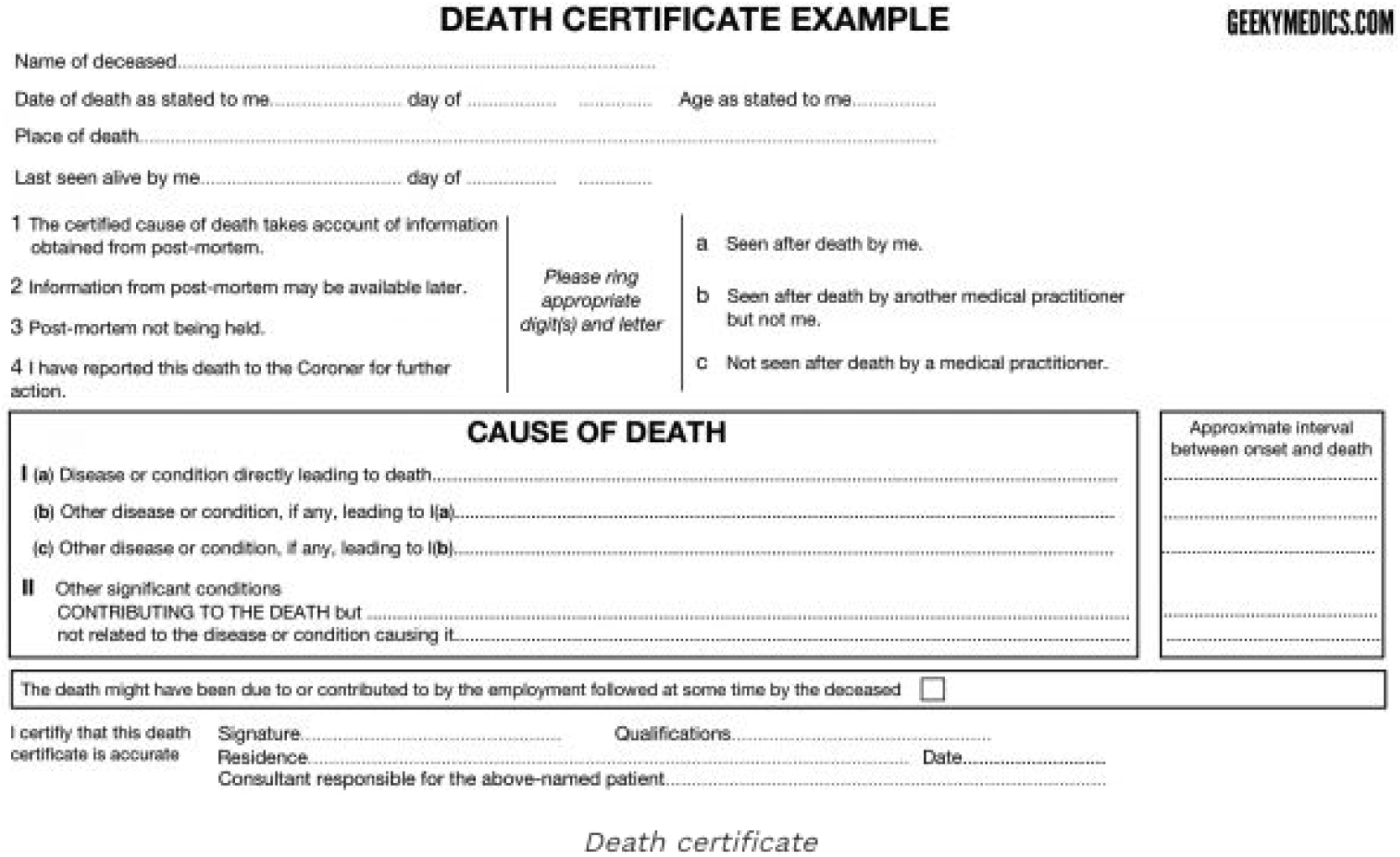

*Facsimile of a death certificate. Image credit:* https://geekymedics.com/certification-death-uk-osce-guide/.

> *‘****Part 1***. *The* ***immediate***, *direct cause of death is reported on the certificate. Then the medical practitioner should go back through the sequence of events or conditions that led to death on subsequent lines until reaching the one that started the fatal sequence. The condition on the lowest completed line of part I will have caused all of the conditions on the lines above it. This initiating condition, on the lowest line of part I will usually be selected as the* ***underlying*** *cause of death, following the ICD coding rules. The WHO defines the* ***underlying*** *cause of death as “a) the disease or injury which initiated the train of morbid events leading directly to death, or b) the circumstances of the accident or violence which produced the fatal injury”. From a public health point of view, preventing this first disease or injury will result in the greatest health gain*.
>
> ***Part 2***. *Other diseases, conditions, or events that contributed to the death, but were not part of the direct sequence, are recorded in part two of the certificate. The conditions mentioned in part two must be known or suspected to have* ***contributed*** *to the death, not merely be other conditions that were present at the time*.*’*

NHS England records the number of patient deaths, therefore, includes **any mention** on the death certificate, whether as an underlying cause or not, and can consist of those who did not test positive for SARS-CoV-2.

#### UK Health Security Agency (UKHSA)

The UKHSA collates reports from multiple sources to provide a daily number of deaths in those with a positive PCR SARS-CoV-2 test or rapid lateral flow test in England, regardless of where they died.

##### Definition

<u>Technical summary</u> UK Health Security Agency data series on deaths in people with COVID-19 (Feb 2022)

> *‘There are* ***two definitions*** *of death in a person with COVID-19 in England, one broader measure and one measure reflecting current trends:*
>
> 1. *A death in a person with a positive SARS-CoV-2 test and either died within* ***60 days*** *of the first specimen date of the most recent episode of infection or died more than 60 days after the first specimen date of the most recent episode of infection, only if COVID-19 is mentioned on the death certificate*.
> 2. *A death in a person with a positive SARS-CoV-2 test and died within (equal to or less than)* ***28 days*** *of the first positive specimen date of the most recent episode of infection*.*’*

#### Care Quality Commission (CQC)

The Care Quality Commission (CQC) publishes statistics on deaths involving COVID-19 in care homes in England.

##### Definition

<u>Transparency statement</u> (2021)

> *‘The inclusion of a death in the published figures as involving COVID-19 is based on the* ***statement of the care home provider***, *which may or may not correspond to a medical diagnosis or test result or be reflected in the death certification. We do not undertake any validation of the data provided to us. We do not contact the provider about the data, except in cases where the form is incomplete*.*’*

#### Northern Ireland

The Northern Ireland Statistics and Research Agency. Cause of Death Information in Northern Ireland: <u>A user guide</u>

##### Definition

The Cause of Death section of the MCCD is set out in two parts as shown below.

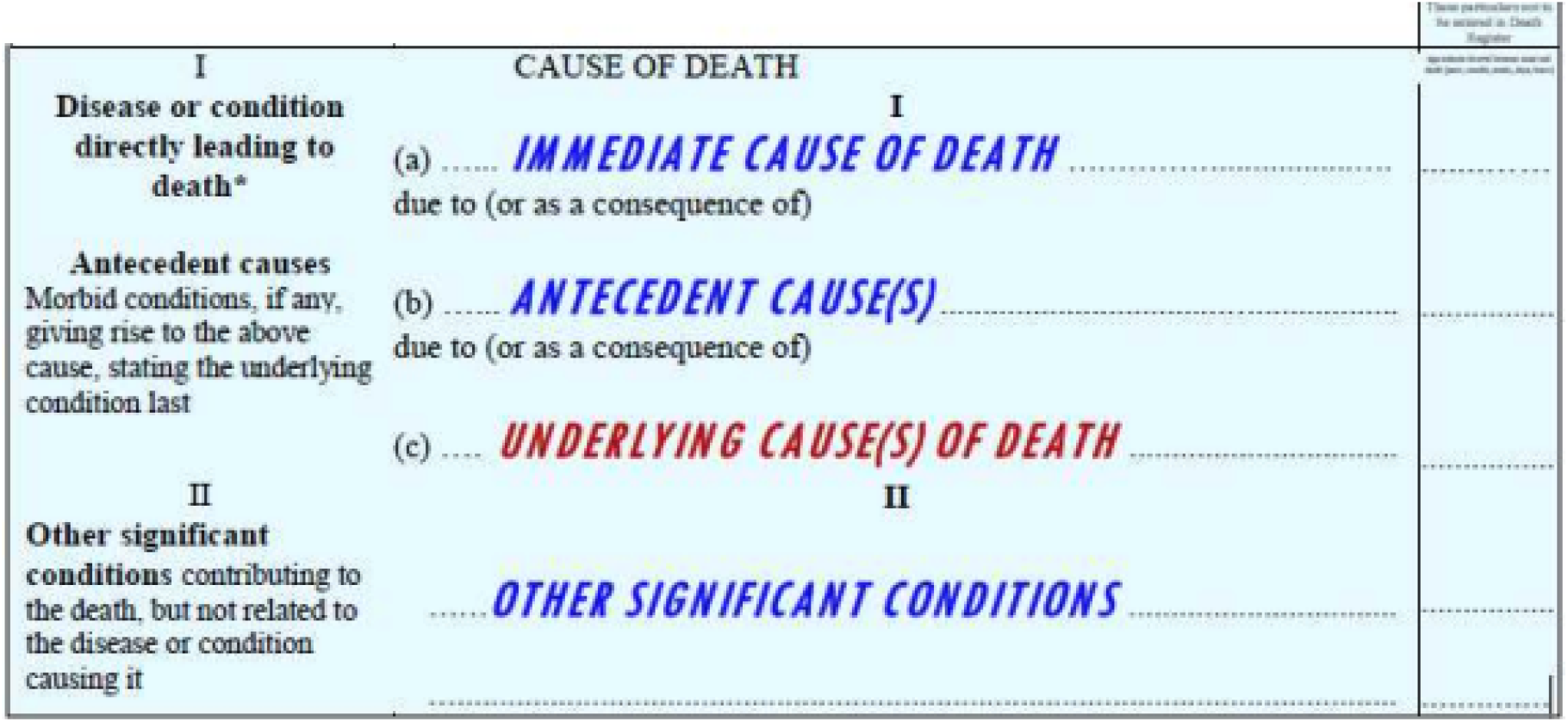

> *‘Part 1 contains the disease or condition directly leading to death and any antecedent causes (i*.*e. conditions that gave rise to the condition leading to death) with the* ***underlying*** *cause stated last*.
>
> *Part 2 contains significant conditions that contributed but were not related to the diseases causing the death*.
>
> *The* ***underlying*** *cause of death is defined as: (a) the disease or injury which initiated the train of morbid events leading directly to death; or (b) the circumstances of the accident or violence which produced the fatal injury*.
>
> *NISRA defined* ***‘sole’*** *or ‘****only****’ cause as the number of cases where ONLY Covid-19 was mentioned on the death certificate and nothing else*.*’*

##### FOI response <u>No 80</u>

> *‘NISRA defines a COVID-19 death as any death where COVID-19 appears anywhere on the death certificate and will include deaths where COVID-19 is the probable cause of death. The Department defines a COVID-19 death as one which occurs within* ***28 days*** *of a positive COVID-19 test, whether or not COVID-19 was the cause of death*.*’*

#### National Records Scotland

##### Definition

Deaths involving coronavirus (COVID-19) in Scotland: Methodology Guide. <u>16 June 2021</u>

> *‘All deaths where COVID-19 was* ***MENTIONED*** *on the death certificate by the doctor who certified the death. This includes cases where the doctor noted that there was suspected or probable coronavirus infection involved in the death*.
>
> *As a result, these weekly totals are likely to be higher than the daily figures - because the daily updates only include those who tested positive for the virus. Using the complete death certificate allows NRS to analyse a lot of information, such as the location of death and what other health conditions contributed to the death*.*’*

#### Scottish Government

##### Definition

Coronavirus (COVID-19) data <u>7 Mar 2022</u>

> *‘Number of people who have died with confirmed COVID-19 (daily deaths)*
>
> *PHS determines confirmed COVID-19 deaths by linking the daily NRS file for newly registered deaths to testing data, and defines a* ***CONFIRMED*** *COVID-19 death as an individual who dies within* ***28 DAYS*** *of their first positive COVID-19 laboratory report (PCR or LFD test), and also within 28 days of possible reinfection, where they have tested positive 90 or more days after their last positive test*.
>
> *This definition was updated on 31 July 2020 to align with the revised definition being used by PHS. This definition was revised on 10 February 2022 to include LFD test results following the change in testing policy. Deaths identified using LFD test results are included in the cumulative total from 6 January onwards. The definition was revised on 1 March 2022 to include possible reinfections. Deaths identified following possible reinfections are included in the cumulative total from the start of the pandemic*.*’*

#### Wales

There are two main sources of COVID-19 mortality statistics in Wales: rapid mortality surveillance data produced by Public Health Wales (PHW) and data by the ONS based on the information collected on the death certificate. See the <u>Technical Advisory Group</u>: Examining Deaths in Wales associated with COVID-19.

##### Definition

> *‘Deaths* ***INVOLVING*** *refer to where COVID-19 was mentioned anywhere on the death certificate as a contributory cause of death. Deaths* ***NOT*** *involving refer to where COVID-19 was not mentioned anywhere on the death certificate as a contributory cause of death*.
>
> *Deaths* ***DUE*** *to COVID-19* ***‘****These refer to where COVID-19 was identified as the* ***UNDERLYING (MAIN)*** *cause of death on the death certificate. Up to January 2021, 89% of registered deaths involving COVID in Wales were due to COVID-19*.*’*

##### Interpretation

There is no consistency in the definition of cause of death or contributory cause of death across national bodies and in different bodies within the same nation.

Medical practitioners and nursing home providers can both assign a cause of death. However, when COVID-19 ONLY is mentioned on the death certificate it is impossible to ascertain a chain of causality. It is also very unlikely that infections with SARS-CoV-2 per se could cause death, in the absence of contributory factors, comorbidities, and a pathology directly deriving from the infection leading to death (such as respiratory failure due to bilateral bronchopneumonia).

Some statements are not clear, and the lack of consistency has confused the public and led to erroneous conclusions. For example, the underlying cause of death is essential for establishing the chain of events. However, there is no clear unified method to establish this vital chain of causation. Further confusion is added because guidance can contradict itself when updates occur. It also seems to be possible to report a death as COVID-19 despite COVID-19 not being the cause of death.

We found 14 different ways to express the cause of death (highlighted above in bold text). The lack of clarity affects the ascertainment and establishment of the causes of COVID-19 deaths and affects the responses reported in sections 2, 3 and 4 of this report.

### 2. Ascertainment of the Cause of COVID-19 Death

#### Background

Several FOIs on the ascertainment of COVID-19 deaths asked about the use of post-mortems (an examination of a body after death), the use of specific tests, or the reporting of different sections on the death certificate.

#### Responses

##### 2a. Of the total deaths, how many have received a post-mortem or equivalent pathological examination to confirm the cause of death?

***Response:* <u>No13</u>**. *NHS England, July 20:*

‘NHS England does not hold the information in relation to the question’

***Response* <u>No84</u>**. *Lothian NHS Board Nov 20:*

‘I am advised by the Mortuary that they do not have access to death certificates, but in the clinical diagnosis section of the Post Mortem report i.e. information provided by the clinician, there were 16 patient cases where Covid-19 or other Covid associated terminologies or phrasing was mentioned.’

##### 2b. How many of the deaths attributed to Covid19 in the UK so far have had a specific test done to determine they have Covid19 and how many have been determined by other means?

***Response:* <u>No17</u>**. *Public Health England, June 20:*

‘The statistics published daily on the PHE dashboard provide the number of deaths of people who have had a positive test for COVID-19. You can find the relevant information at the following <u>link</u>’

##### 2c. I would like to know on which rows/sections of the death certificate there was a mention of COVID, if any, for the totals presented each week in the row 14 data?

***Response:* <u>No19</u>**. *Office for National Statistics, May 20*:

‘I am writing to confirm that the Office for National Statistics has now completed its search for the information which you requested and the response can be found <u>here</u>’

‘Some of the analysis requested is available through different releases. On 16 April 2020, we released an Analysis of deaths involving COVID19, which stated that, including registrations up to 6 April, there were 3,912 deaths involving the coronavirus (COVID-19) that occurred in March 2020 in England and Wales; of these, 3,372 (86%) had COVID-19 assigned as the underlying cause of death.

This analysis will be updated on 15 May to include deaths occurring in April.

The number of deaths involving COVID-19 by where it is mentioned on the death certificate is available to commission as a bespoke analysis. Such services would fall outside of the Freedom of Information regime and would be subject to legal frameworks, disclosure control, resources and agreement of <u>costs</u>, where appropriate.’

##### 2d. Please provide the criteria by which death by COVID-19 is distinguished from death by influenza, pneumonia, asthma, and other respiratory illnesses

***Response* <u>No32</u>**. *University Hospitals of Derby and Burton NHS Foundation Trust*

‘Most clinicians would ascribe a death to COVID if a) there was a positive swab in the lead up to the death, or b) the clinical pattern of disease is such that, despite a negative swab, COVID was the most likely cause’

#### Interpretation

The dearth of post-mortem examinations, added to the lack of consistency in the definition of what is meant by a COVID-19 death and inconsistent testing, enhances the difficulty to ascertain the direct cause of death.

Guidance for doctors completing Medical Certificates of Cause of Death in England and Wales FOR USE DURING THE EMERGENCY PERIOD ONLY

Deaths are required by law to be registered within 5 days of their occurrence unless there is to be a coroner’s post mortem or an inquest. If no doctor attended the deceased within 28 days of death or the deceased was not seen after death by a doctor, the MCCD can still be completed. However, the doctor will be obliged to refer the death to the coroner before it can be registered. The coroner may instruct the registrar to accept the certifying doctor’s MCCD for registration.

Where a cause of death cannot be ascertained, the death cannot be certified, and the doctor should refer the death directly to the coroner with any supporting information. This information will be used for mortality statistics, but the death will be legally “uncertified” if the coroner does not investigate and make a determination as to the cause of death.

Strictly speaking, the law requires that an MCCD should be completed even when a death has been referred to the coroner. In practice, if the coroner has decided to order a post-mortem examination and/or to hold an inquest, he may tell a doctor not to complete the MCCD. However, the coroner can only legally certify the cause of death if he has investigated it through autopsy, inquest or both. This means that, if the coroner decides not to investigate, the registrar will need to obtain an MCCD from a doctor who attended the deceased before the death can be registered.

*Publication:* <u>Guidance for Doctors</u>

### 3. COVID-19 as the sole cause of death or died with no underlying health issues

#### Background

The ‘sole cause of death’ FOIs relates to the underlying health and attempts to understand those that died with no other related health conditions. Underlying health conditions also includes FOIs on the number of do not resuscitate orders that were in place at the time of death.

#### Responses

##### 3a. What is the actual number of deaths where the sole cause of death was COVID-19 with no underlying medical health issues?

***Response* <u>No21</u>**. *UK Statistics Authority, May 20:*

‘I am writing to confirm that the Office for National Statistics has now completed its search for the information which you requested and the response can be found <u>here</u>:’

***Response* <u>No30</u>**. *UK Statistics Authority, Nov 20:*

‘We are no longer publishing monthly COVID-19 deaths with details around the death such as pre-existing conditions or no underlying cause. This is due to the decrease in the number of COVID-19 related deaths. Whilst the number of infections appear to be rising, the number of COVID-19 deaths from July onwards are very small and pose a disclosure risk, making it possible to identify individual records.’

##### 3b. I would like to know how many COVID 19 ONLY deaths have been registered in 2020, in Scotland. NOT COVID 19 related deaths

***Response* <u>No113</u>**. *National Records of Scotland, Jan 21:*

‘In our weekly Covid-19 statistics publication, we provide figures on both the number of deaths where Covid-19 was mentioned on the death certificate (either as the underlying cause or as a contributory factor) and on deaths where Covid-19 was the underlying cause of death. For this reason, the information you require for deaths where Covid-19 was the underlying cause of death is available from Table 3 (2021) and Table 3 (2020) in the weekly publication Deaths involving coronavirus (COVID-19) in Scotland | National Records of Scotland (nrscotland.gov.uk) the most recent of which can be found <u>here</u>’

##### 3c. How many deaths in the NHS trust where COVID-19 was the ONLY cause of death?

***Response* <u>No122</u>**. *Royal Cornwall Hospitals NHS Trust, Mar 21:*

‘10 - From the 01st January 2020 to 31st January 2021 10 patients have died across the Royal Cornwall Hospitals Trust where Covid-19 alone has been written on the death certificate. That means no other cause of death at all was detailed on the Medical Certificate of Cause of Death written by the doctor’

***Response* <u>No137</u>**. *Oxford Health NHS Foundation Trust, Jan 21:*

‘To confirm, there was only 1 death during the period (2020) where Covid 19 was the only cause of death, in April 2020, of a person who was 82 years old.’

***Response* <u>No138</u>**. *North Tees and Hartlepool NHS Foundation Trust, Mar 21:*

‘Furthermore, patients may have ‘solely’ died of Covid-19 in terms of what has been determined as their cause of death, however, the Trust does not undertake post mortems and therefore we are unable to say whether they had underlying, undiagnosed co-morbidities which contributed to their death.

The Trust is following national government guidance which states that anyone who has died within 28 days of a positive Covid-19 test is classified as dying with Covid-19 and included in our figures which are returned to the national team. The Government/NHS England may have further information that is not kept or recorded at a Trust Level.’

***Response* <u>No141</u>**. *Ayrshire and Arran NHS Board, Feb 21:*

‘for the specified time period, there have been 338 deaths where COVID-19 is mentioned on the death certificate, with 292 where COVID-19 is cited as the primary cause of death. Of these deaths, there have been 11 where no other conditions are mentioned apart from COVID-19.’

***Response* <u>No142</u>**. *Grampian Health Board, Apr 21:*

‘There were <5 individuals whose sole cause of death was Covid-19 at Aberdeen Royal Infirmary in that time period. Due to the very small number involved the exact figure cannot be disclosed as this may identify individual patients.’

***Response* <u>No146</u>**. *Cwm Taf Morgannwg University Health Board, Jan 21:*

‘Nine - For the date period 01/02/2020 – 31/12/2020 There have been 9 deaths in Cwm

Taf Morgannwg University Health Boards Princess of Wales Hospital only, with no comorbidities.’

***Response* <u>No149</u>**. *Cwm Taf Morgannwg University Health Board, Jan 21:*

‘78 - For the date period 01/02/2020 – 31/12/2020 There have been 78 resident deaths in the Cwm Taf Morgannwg University Health Board area, with no comorbidities.’

***Response* <u>No156</u>**. *University Hospitals of Leicester NHS Trust, Mar 21:*

‘As of 16:00 hours on 15 March 2021, the University Hospitals of Leicester NHS Trust had reported 1,187 patient deaths from 1 February 2020 up to, but not including 17 February 2021, where COVID-19 was documented as the direct cause of death on part 1 of the death certification process.’

***Response* <u>No110</u>**. *South Tees Hospitals NHS Foundation Trust, Feb 21*

‘The Trust has had 32 deaths with no underlying health conditions.’

##### 3d. Of the deaths reported as being caused by COVID-19, how many had underlying health issues?

***Response:* <u>No28</u>**. *NHS England, Apr 20:*

‘NHS England does not hold this information.’

##### 3e. Part 1. Can you explain how every single person who has pre-existing conditions has died within 28 days of a positive test and every single person who is labelled as having no pre-existing conditions has not had a positive test in the last 28 days?

***Response* <u>No65</u>. *part 1***. *NHS England Jan 21:*

‘NHS England publishes the number of COVID-19 deaths that occurred in hospitals in England and whether they had a pre-existing condition or not in the weekly file on our <u>website</u>

The weekly data published on 10 December 2020 showed 43,537 deaths of patients who have died in hospitals in England and have tested positive for COVID-19. The daily total deaths publication on the same day showed (separately) a further 2,092 deaths where a positive test result for COVID-19 was not received but COVID-19 is mentioned on their death certificate.

The weekly data set shows on Table 3 that of the 43,537 patients that had tested positive for COVID-19, 41,683 patients had one or more pre-existing conditions and 1,854 patients had no pre-existing condition. We do not publish data on pre-existing conditions for the patients with no positive test result for COVID-19.’

##### 3e. Part 2. If a person is diagnosed as dying from Covid 19 but has no ‘pre-recorded’ pre-existing conditions and the rules for the Medical Certification for Cause of Death have been changed to allow Cause of Death to be ruled in favour of Covid 19, this then allows for a huge margin of error from fact

***Response* <u>No65</u>. *part 2***. *NHS England, Jan 21:*

‘The pre-existing conditions reported to NHS England are not necessarily the same as what was noted on a patient’s death certificate.’

##### 3f. Can you please tell me the number of people who have died from Covid 19 without any pre-existing medical conditions?

***Response* <u>No87</u>**. *Mersey Care NHS Foundation Trust, Dec 20:*

‘The number of patients who have died of Covid-19 is already in the public domain: (see <u>here</u>). All patients who have died within Mersey Care had pre-existing conditions.’

##### 3g. How many patients had DNACPR (Do Not Attempt Cardiopulmonary Resuscitation Order on their records?

***Response* <u>No65</u>**. *London North West University Healthcare NHS Trust, Dec 20:*

‘555 (*of 623 deceased patients who tested positive for COVID-19)

*Please note that although these 555 (*DNACPR, Do Not Attempt Cardiopulmonary Resuscitation Order) patients tested positive for COVID-19, not all 555 were noted to have died as a direct result of COVID-19. To confirm, 138 had COVID-19 recorded as a contributing factor in the patients cause of death, with 417 having COVID-19 recorded as the main cause of death (*Part 1a of Death Certification)’

***Response* <u>No77</u>**. *Barts Health NHS Trust, Nov 20:*

‘How many of these 649 patients had DNACPR orders (or similar decisions) on their records? 207 patients that had died with COVID within our hospitals as of 4pm on 11 October 2020 had DNACPR orders in place.*’*

***Response* <u>No85</u>**. *Liverpool University Hospitals NHS Foundation Trust. Nov 20:*

*‘(*from April 2020 to October 2020) Royal & Broadgreen data: 353 COVID related deaths were reported for Royal Site. Of these, 337 had a DNAR in place.’

***Response* <u>No89</u>**. *Pennine Acute Hospitals NHS Trust, Oct 20:*

‘A review has been undertaken for all deaths where COVID was recorded between July to September 2020. A total of 43 COVID related deaths occurred at the Trust in this time period. However, 33 records were available for review at the time of this request. Of the 33 records reviewed, 30 had a DNACPR form in place at the time of death.’

***Response* <u>No93</u>**. *King’s College Hospital NHS Foundation Trust, Nov 20:*

‘How many of these 515 patients had DNACPR orders (or similar decisions) on their records? 440’

***Response* <u>No116</u>**. *Betsi Cadwaladr University Health Board, Feb 21:*

‘The Health Board can confirm that we do record do not attempt resuscitation (DNAR) status. However, in order to gather this information, we would have to carry out a specific exercise of reviewing all the Medical Certificate Cause of Death (MCCD) books for each patient in each hospital to establish whether COVID-19 was in present and then check DNAR against our records. We have established that to comply with your request would exceed the appropriate costs limit under Section 12 of the Freedom of Information Act 2000.’

#### Interpretation

Identification of the main cause of death is vital to assessing the severity of the pandemic and permitting meaningful comparisons of death data. The already noted vagueness of the meaning of testing, as well as inconsistencies in definition use, undermine assessments of the role of infection with SARS-CoV-2 and cause of death in relation to other underlying health conditions.

COVID-19 as t<u>he sole cause of death</u> is uncommon in frail home healthcare individuals in Sweden; however, it isn’t uncommon on death certificates in the UK (see <u>NISRA</u>).

ONS produce a quarterly report on the pre-existing conditions of people who died due to COVID-19, in England and Wales (see <u>here)</u>.^12^ In December 2021, ONS <u>published an FOI response</u> asking about deaths from COVID-19 with no other underlying causes, and a <u>blog post</u> in January 2022 that reported claims that only 17,000 people died from COVID-19 was ‘highly misleading’.^13^

Whether or not the deceased had a do-not-resuscitate order (DNR) is not recorded on the death certificate. In the trusts reporting this number, it was high. For example, in the North-West University Healthcare NHS Trust 89% of 623 deceased patients positive for COVID-19 had DNACPR in place, and in the King’s College Hospital NHS Foundation Trust the proportion was 85%. However, in the Barts Health NHS Trust, the proportion was lower at 32%. This variation warrants further investigation.

In an FOI response to deaths due to COVID-19 with no pre-existing conditions by county/city, the <u>ONS produced a table</u> of the counts of deaths registered in England and Wales by the Local Authority of residence and place of death.^14^

The results for 2020 report that in care homes in England 1,304 out of 17,264 COVID-19 deaths were registered with no pre-existing conditions (unweighted average 7.6%, range 0% to 63%). See https://datawrapper.dwcdn.net/5Bo8A/4/ for the analysis by local authorities.

The lack of pre-existing conditions in care home residents (on average one in thirteen and in some homes more than half of the COVID-19 deaths had no pre-existing conditions at the time of death) further adds to the uncertainties over the assignment of causation. Care home residents have multiple comorbidities that contribute to their vulnerability, need for additional care, and usually their death.

The assignment of death with no pre-existing conditions, therefore, seems implausible and may follow on from the vagueness of the guidelines for care home certification <u>provided by the CQC</u>, whereby death involving COVID-19 can be based on the statement of the care home provider.

### 4. Died WITH or FROM COVID-19

#### Background

FOI questions in this section aim to determine who is susceptible to dying because of pre-existing conditions and therefore died ‘with’ COVID versus those that died as a result - “from” COVID.

#### Responses

##### 4a. Please provide the recorded figures per month since January 2020, per hospital, for deaths caused by as opposed to died WITH COVID-19

***Response* <u>No31</u>**. *University Hospitals of Leicester NHS Trust, May 21:*

‘This is distinguished by the Medical Examiner (an independent doctor who provides scrutiny on all deaths) having a conversation with the doctor issuing the Medical Certificate of Cause of Death (MCCD). This conversation focuses on the reasons for admission, clinical examination findings, findings from relevant investigations such as swab results, viral tests, chest X-ray, full blood count, C-reactive protein result and CT chest findings, and the working diagnosis from the clinical team looking after the patient. Covid-19 has a characteristic clinical pattern and when this is present with a positive swab result, and in the absence of positivity for other viruses, a diagnosis of Covid-19 would usually be made.

The Medical Examiner and doctor would also discuss what contribution Covid-19 has made to the death. Sometimes it is clear it is the cause, sometimes it is felt it is more of a contributing cause in which case it is put into part 2 of the Medical Certificate of Cause of Death. Sometimes it might be felt that Covid-19 did not contribute at all (e.g. in a patient who is dying of terminal cancer) and therefore it may be omitted from the death certificate despite a positive test result. All of this process relies on the expertise, training and judgement of the doctors involved and is tailored to the unique circumstances of each patient.’

***Response* <u>No32</u>**. *University Hospitals Derby & Burton NHS Foundation Trust, Dec 20:*

‘Most clinicians would ascribe a death to COVID if a) there was a positive swab in the lead up to the death, or b) the clinical pattern of disease is such that, despite a negative swab, COVID was the most likely cause.’

***Response* <u>No33</u>**. *Nottingham University Hospitals NHS Trust, Dec 20:*

‘I can confirm that the department holds information that you have asked for. The information is exempt under section 21 of the FOI Act because it is reasonably accessible to you, and I am pleased to inform you that you can access it via the following <u>link</u>’

***Response* <u>No34</u>**. *Liverpool University Hospitals NHS Foundation Trust, Jan 21:*

‘Please be advised in relation to your questions above we cannot provide data relating to the “sole diagnosis” of a patient. Neither can we provide the “direct cause” of a patient’s death as we do not have access to patient’s death certificates.’

##### 4b. What is the total number of deaths ‘with Covid’ and what is the total number of deaths ‘from Covid’?

***Response:* <u>No37</u>** *York Teaching Hospital NHS Foundation Trust, Mar 21:*

‘Any COVID death 575 (These deaths are people who have died in hospital and have had a positive). Deaths “from” COVID Covid test – we can not determine whether or not Covid was the cause of death, i.e. death ‘from’ Covid.’

##### 4c. Please state what criteria are used to designate death as having been due to the above viruses for statistical purposes?

***Response:* <u>No43</u>**. *Northern Ireland Statistics and Research Agency, Jan 21:*

‘NISRA, along with the other UK jurisdictions recognised the need for a mentions based approach to Covid-19 deaths reporting given the coverage limitations of the Department of Health-related, positive test based statistics which at the start of the pandemic largely excluded deaths outside of the hospital; and also given that statistics based on Covid-19 as the underlying cause of death are not available in NI until some time after each quarter ends to allow for ICD-10 coding to take place.

Figures on this basis are currently available up to the end of September 2020 at; https://www.nisra.gov.uk/publications/registrar-general-quarterly-tables-2020‘

##### 4d. Evidence that the ‘COVID-19’ related deaths are in fact deaths from the virus solely and not just deaths recorded with the smallest hint of being around COVID-19, a cough, temperature, etc

***Response* <u>No49</u>**. *Welsh Government, Jan 21:*

‘The Office for National Statistics (ONS) collect mortality data for England and Wales based on information collected on the death certificate. The ONS publish the number of reported deaths by method of certification and registration (including the number of post mortems) for England and Wales in their user guide, but this is currently unavailable for the pandemic period.’

##### 4e. I require the number of deaths from COVID 19 alone, i.**e. patients dying OF COVID 19, not WITH it, having no other comorbidity**

***Response* <u>No54</u>**. *East Lancashire Hospitals NHS Trust, Dec 20:*

‘Number of Deaths where COVID-19 / Coronavirus is noted in Part 1 of the Death Certificate – 287 (01/04/2020 - 25/11/2020)’

***Response* <u>No57</u>**. *Medway NHS Foundation Trust, Nov 20:*

‘267 deaths February 2020 and October 2020’

***Response* <u>No108</u>**. *PHE, Feb 21:*

‘PHE can confirm it holds this information which is exempt under Section 21 of the Act as it is reasonably accessible by other means. For your convenience, I have included a link below to the Coronavirus dashboard which displays deaths with COVID-19 mentioned on the death certificate: https://coronavirus.data.gov.uk/details/deaths‘

***Response* <u>No147</u>**. *North West Anglia NHS Foundation Trust, Feb 21:*

(f) Of the 171 deaths recorded at Hinchingbrooke Hospital between 20/03/2020-27/01/2021, we cannot accurately state how many patients died with COVID-19 as the underlying cause of death. Please note that the number of deaths reported includes patients whose most recent swab was positive prior to their death, as well as those patients whose most recent swab, was negative but COVID-19 was listed on their death certificate.

#### Interpretation

The inconsistencies already noted hinder the ascertainment of the role of each factor leading to death and the quantification of the importance of infection. Some responses indicate that SARS-CoV-2 negative individuals were classified as COVID-19 deaths.

## Discussion

The Victorian physicians John Snow and his aide Duncan laid the epidemiological groundwork in the ascertainment of death during an ugly cholera outbreak in central London. Snow and Duncan enquired door to door as to whether deaths had occurred, when and where.

They matched death certificates with the results of their enquiries as they realised that ill people could have moved away from Soho and died elsewhere. By this systematic approach, Snow was able to group like with like (all death caused by cholera), narrow down a possible source (contaminated water) and mode of transmission (waterborne). However, he was also able to define the severity of the outbreak. Although cholera has very characteristic manifestations its transmission is not 100% linear and Snow did not have the benefit of an identified agent, electron microscopy or a modern system of testing. The lack of rigorous approach to cause of death definition and attribution are possibly due to the fog of the pandemic but also point to lack of preparation and lack of control and supervision of the events from 2020.

Observed differences, whether between agencies or between UK countries are affected by the definitions used. Statistical comparisons with other countries should also be treated with caution as they will also be substantially affected by the definitions chosen.

The FOI question of ‘**with**’ or ‘**for**’ COVID-19 death remains a central issue to understand the impact of the pandemic. This question cannot be answered with any certainty through the sole use of death certificates, particularly given their inherent limitations.

In hospitals, junior doctors can often be <u>tasked with</u> signing medical certificates of cause of death.^15^ Normally, the doctor who verifies the death, or cared for the patients during their last 14 days is eligible to sign the MCCD. In the pandemic, the duration was <u>extended to 28 days</u>.

A death certificate is based on the probability the deceased expired of the causes on the death certificate. Prioritising the condition leading to death can prove difficult: it is affected by the experience of the clinician, their prior knowledge of the patient. In the presence of several comorbidities that may compete and co-exist with each other an individual may easily appear to die with rather than of their disease.^16^

The condition listed on the bottom line of Part I of the certificate is the underlying cause of death. The cause of death is based on medical opinion, which may change as more information becomes available.

In the midst of a pandemic, the assignment of causation will be affected by availability and representativeness heuristics. Heuristics are mental shortcuts that aid problem-solving and judgments. However, they can often lead to erroneous conclusions.^17^

The availability heuristic also referred to as the <u>availability bias</u> is ‘a distortion that arises from the use of information which is most readily available, rather than that which is necessarily most representative.’^18^

Determining the underlying cause of COVID-19 deaths remains an important area of future research. The high rates of Do Not Resuscitate orders in some Trusts for COVID-19 deaths and the variation in rates requires further investigation. Were these orders already in place, as may be the case for those with a terminal disease, or were they instigated during admission? These vital questions require answering - to determine the impact of the virus, particularly in those that are the most vulnerable and most likely to die.

The current FOIA prevents vital responses due to the limitations on the time required to assess some of the answers. We consider an ombudsman should override the time allowance when the answer is in the public interest and will facilitate public understanding.

The assignment of the causation of deaths can be done using several methods. The anonymised death certificate data set with the conditions and their specific placement on the death certificate would permit independent analysis of the assignment of the underlying cause of death.

Confirmation of the cause of death could be established by the use of post-mortem evidence in the presence of the full documentation of the medical history of the deceased. A subset of deaths could be verified using autopsies with full medical documentation. However, the UK guidelines and the FOI answers we reviewed suggest autopsies were an uncommon practice.

An FOI <u>response by the Welsh Government</u> stated ‘the ONS publish the number of reported deaths by method of certification and registration (including the number of post-mortems) for England and Wales in their user guide, but this is currently unavailable for the pandemic period.’ An <u>FOI request to the ONS</u> asked about the number of autopsies carried out. The response stated, the ONS ‘do not hold analysis on the number of post-mortems completed.’

The ONS can create an analysis on deaths involving post-mortems in 2020 and 2021 subject to legal frameworks, disclosure controls, resources, and agreement of costs. The number of post-mortems carried out in the UK is currently unclear.

As we go down the causality pyramid (see figure) the uncertainty over the true cause of death increases.

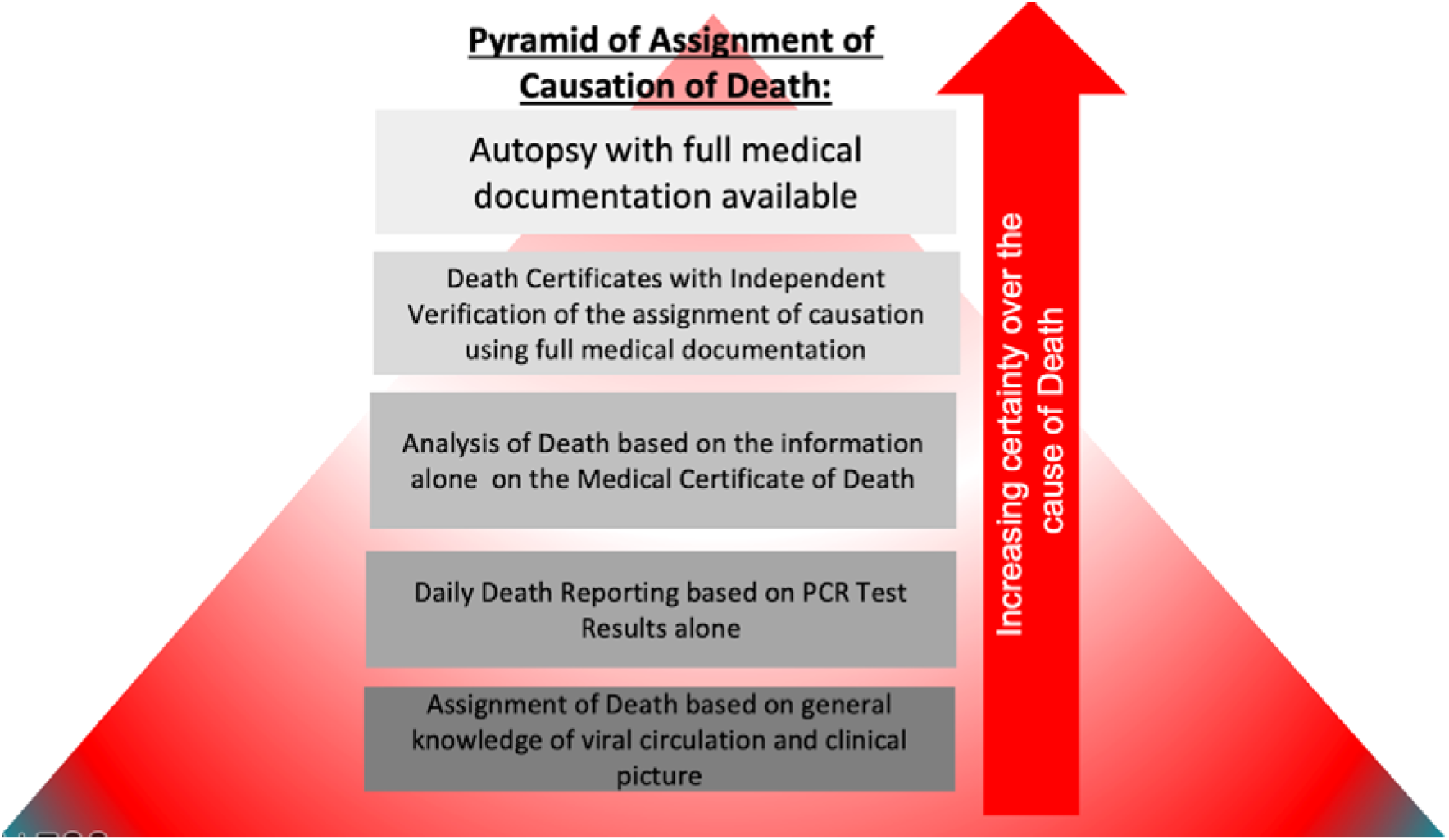

Analysis of a random sample of death certificates assigned as ‘**from**’, ‘**with**’, ‘**underlying**’, ‘**due to**’, or ‘**involving**’ COVID-19 and exploration of their underlying causes with independent verification of the assignment of causation using full medical documentation would reduce uncertainty. One level down is based on reporting of a positive test (usually a PCR with no further detail). At the bottom of the pyramid is Daily Death Reporting based on knowledge of the pandemic on general knowledge of viral circulation and the clinical picture.

## Conclusion

The current system is seeding confusion in the public (as shown by the number of questions asked) and damaging trust in the assignment of the cause of the COVID-19 deaths. A coherent process based on consistent definitions across the devolved nations is required. Furthermore, the system requires validation through the addition of post-mortem data when possible to ensure accuracy.

## Data Availability

All data included in the review are available via Google Docs.

https://docs.google.com/spreadsheets/d/1MU7efHDkJETZgUYCn1lwnzRf9hn2baOC/edit?usp=sharing&ouid=100001257459010812550&rtpof=true&sd=true

